# HIV and SARS-CoV-2 infection in postpartum Kenyan women and their infants

**DOI:** 10.1101/2022.06.23.22276827

**Authors:** Emily R. Begnel, Bhavna H. Chohan, Ednah Ojee, Judith Adhiambo, Prestone Owiti, Vincent Ogweno, LaRinda A. Holland, Carolyn S. Fish, Barbra A. Richardson, Adam K. Khan, Rabia Maqsood, Efrem S. Lim, Dara A. Lehman, Jennifer Slyker, John Kinuthia, Dalton Wamalwa, Soren Gantt

## Abstract

**Background:** HIV may increase SARS-CoV-2 infection risk and COVID-19 severity generally, but data are limited about its impact on postpartum women and their infants. As such, we characterized SARS-CoV-2 infection among mother-infant pairs in Nairobi, Kenya.

**Methods:** We conducted a nested study of 53 HIV-uninfected and 51 healthy women living with HIV, as well as their HIV-exposed uninfected (N=41) and HIV-unexposed (N=48) infants, participating in a prospective cohort. SARS-CoV-2 serology was performed on plasma collected between 1 May-31 December 2020 to determine the incidence, risk factors, and symptoms of infection. SARS-CoV-2 RNA PCR and sequencing was also performed on stool samples from seropositive participants.

**Results:** SARS-CoV-2 seropositivity was found in 38% of the 104 mothers and in 17% of the 89 infants. There was no significant association between SARS-CoV-2 infection and maternal HIV (Hazard Ratio [HR]=1.51, 95% CI: 0.780-2.94) or infant HIV exposure (HR=1.48, 95% CI: 0.537-4.09). Maternal SARS-CoV-2 was associated with a >10-fold increased risk of infant infection (HR=10.3, 95% CI: 2.89-36.8). Twenty percent of participants had symptoms, but no participant experienced severe COVID-19 or death. Seroreversion occurred in ∼30% of mothers and infants. SARS-CoV-2 sequences obtained from stool were related to contemporaneously circulating variants.

**Conclusions:** These data indicate that postpartum Kenyan women and their infants were at high risk for SARS-CoV-2 infection in 2020, and that antibody responses waned rapidly. However, most cases were asymptomatic and healthy women living with HIV did not have a substantially increased risk of infection or severe COVID-19.

## INTRODUCTION

To date, SARS-CoV-2 has infected >246,000 people and caused >5,000 deaths in Kenya [1]. Kenya’s first COVID-19 case was reported on March 12, 2020 [2] and generalized community spread was recognized by May, which was followed by two major waves of infections between July-September and October-December 2020 [1,3]. Surveillance suggests most diagnosed SARS-CoV-2 infections in Kenya at this time were asymptomatic [1]. Through 2021, Kenya has experienced several subsequent waves with higher numbers of hospitalizations and deaths [1,3], concurrent to emergence of several variants of concern [4].

Kenya has an estimated 1.5 million adults and children living with HIV [5]. Persistent inflammation and immune dysregulation are hallmarks of HIV that are only partially resolved by suppressive antiretroviral therapy (ART) [6], and can result in pulmonary, cardiac, and other comorbidities that are risk factors for severe COVID-19 [7]. Several studies and recent meta-analyses suggest people living with HIV may have an increased risk of COVID-19 mortality, which is higher among those not on ART [8–15]. More research is needed to understand whether COVID-19 risk remains elevated in individuals with effectively managed HIV infection.

Little is currently known about the risks and outcomes of SARS-CoV-2 infection among postpartum women or HIV-exposed uninfected (HEU) infants. Immunologic changes postpartum may increase susceptibility to SARS-CoV-2 infection or severity of COVID-19. Additionally, HEU infants, which have a nearly two-fold higher risk of overall mortality than HIV-unexposed uninfected infants (HUU) [16–18], may also be at higher risk of severe COVID-19. However, existing data are limited to the effects of SARS-CoV-2 infection during pregnancy or the neonatal period, and there is a lack of comprehensive data among postpartum women living with HIV and HEU infants.

To address these gaps, we assessed the incidence, risk factors, and symptomatology of SARS-CoV-2 infection among postpartum women, both living with HIV and HIV-uninfected, and their infants who were participating in a longitudinal cohort study in Nairobi, Kenya during the country’s first two waves of COVID-19 in 2020.

## METHODS

Human subjects approvals for all study procedures were obtained from the Kenyatta National Hospital-University of Nairobi Ethics and Research Committee and the University of Washington Institutional Review Board. All participants provided an initial written informed consent for participation in the parent cohort study; an additional written informed consent was required for SARS-CoV-2 serology testing.

### Participants and Follow-up

This study was nested into the Linda Kizazi Study, a prospective cohort study of the infant virome. Between December 2018-March 2020, 211 pregnant women in their third trimester were recruited from Mathare North Health Centre in Nairobi. Women were eligible if aged 18-40 years, between 28-42 weeks gestation, planning to breastfeed, and, if living with HIV, had received ≥6 months of ART. Exclusion criteria included planned Caesarean section, serious medical condition, and taking antimicrobial or immunosuppressive medication other than for HIV prophylaxis.

All mother-infant pairs were followed from delivery through two years postpartum with clinic visits at week 6, week 10, month 6, and every three months thereafter. At each visit, staff collected information about current and recent symptoms of illness, healthcare visits, diagnoses, medications, and immunizations. A physical exam was performed and samples, including blood and stool, were collected. Data on mothers’ sociodemographic characteristics and health and obstetric history were collected at enrollment. For women living with HIV, CD4 testing was conducted at enrollment and every six months postpartum.

### SARS-Co-V-2 Antibody Assays

Plasma samples collected between January 1-December 31, 2020 were tested retrospectively for SARS-CoV-2 antibodies, and samples collected between January 1-March 3, 2021 were tested for all participants with ≥1 positive test in 2020 to evaluate antibody loss. Samples were tested for detection of total antibodies (IgM/IgA/IgG) to a recombinant SARS-CoV-2 nucleocapsid protein using the Platelia SARS-CoV-2 Total Antibody ELISA (Bio-Rad, Marnes-la-Coquette, France), which had US FDA Emergency Use Authorization at study commencement with reported specificity of 94.9% and a sensitivity of 97.4% [19]. We enrolled in an external quality assessment program with the European Society for External Quality Assessment (Heidelberg, Germany) and successfully passed two cycles of proficiency test panels.

The final ELISA result was based on the ratio of the optical density (OD) value of the sample to the mean OD value of the cut-off controls. The result was considered negative for SARS-CoV-2 antibodies if the OD ratio was <0.8, positive if ≥1.0, and equivocal if ≥0.8 and <1.0, according to the manufacturer’s instructions. Samples with equivocal results were retested before final interpretation. Antibody loss was defined by ≥1 negative ELISA result after an initial positive test. For longitudinal modeling of antibody decline, all OD values below the limit of detection (OD ratio=0.8) were set to 0.4, the midpoint between the lower limit and zero.

### Detection and Sequencing of SARS-CoV-2 RNA in Stool

Stool samples collected from seropositive participants on or between their last seronegative and first seropositive time points were tested for viral RNA. Detailed methods for stool viral RNA extraction, RT-PCR, and sequencing are provided in the S1 Appendix. Briefly, total nucleic acid was extracted from homogenized and filtered stool specimens and quantitative real time PCR (qRT-PCR) was performed using the QuantStudio 3 Real-Time system (Applied Biosystems) [20]. Full-length SARS-CoV-2 genome sequencing was attempted on all qRT-PCR-positive samples. Consensus sequences were called using iVar (version 1.0; parameters -q 20, -t 0.75, -m 20, -n N) [21]. Lineages were assigned using pangolin (version 2.3.8) [22]. Sequence alignments were performed with MAFFT (version 7.471) [23] and phylogenetic reconstruction performed with iqtree with 1000 ultrafast bootstraps [24]. Phylogeny was visualized using FigTree (version 1.4.4) [25]. Sequences used in phylogenetic analysis include 500 randomly selected global sequences from GISAID [26], the Wuhan1 reference genome, and the two genomes sequenced in this study (GISAID accession numbers EPI_ISL_2771497 and EPI_ISL_2771498).

### Statistical Analyses

Kaplan-Meier survival analysis was used to estimate the incidence of SARS-CoV-2 infection among mothers and infants separately. Incidence rates (IRs) were calculated as the number of first positive antibody tests per 1000 person-days at risk. Mothers’ time at risk was set to begin on May 1, 2020, when generalized community transmission began in Kenya [1], and infants’ time at risk began either on May 1, 2020 or their date of birth if born later. All participants’ time at risk ended at the estimated time of SARS-CoV-2 infection, date of last negative serology test, or was censored on December 31, 2020. Time of infection was estimated as the midpoint between a participant’s last negative serology test or May 1, 2020 (whichever was later) and their first positive serology test. The log-rank test was used to compare time to infection between women living with HIV and HIV-uninfected women, HEU and HUU infants, and infants whose mother was ever versus never SARS-CoV-2 seropositive. No infants acquired HIV infection.

Cox proportional hazards regression models were used to assess correlates of SARS-CoV-2 infection, both overall and stratified by HIV status/exposure. Growth measures were obtained at infants’ last study visit before May 1, 2020 and include continuous weight-for-age (WAZ), height-for-age (HAZ), and weight-for-height (WHZ) z-scores and the corresponding outcomes of underweight (WAZ <-2), stunting (HAZ <-2), and wasting (WHZ <-2). Z-scores were calculated using the -zanthro-command in Stata using the WHO standard reference and adjusting for sex.

Generalized estimating equations with the log link and an independent correlation structure were used to evaluate the relative risk (RR) of symptoms of COVID-19 associated with participants’ first positive serology result. The outcome was defined as report of ≥1 symptom of COVID-19 experienced either at the time of the visit or since their most recent prior visit (typically a three-month window). Symptoms included those listed by the US Centers for Disease Control and Prevention (CDC), including fever or chills, cough, shortness of breath or difficulty breathing, fatigue, muscle or body aches, headache, new loss of taste or smell, sore throat, congestion or runny nose, nausea or vomiting, and/or diarrhea [27]. Data from both clinic-based and home-based (if applicable) visits were included. All time points prior to the first positive serology test were considered SARS-CoV-2 negative visits for calculation of RRs.

All analyses were conducted in Stata (version 14; StataCorp, College Station, TX, USA) using two-sided tests with a significance level of α=0.05.

## RESULTS

### Participant Characteristics

There were 104 mothers and 89 infants tested for SARS-CoV-2 antibodies (Table 1). Median age at enrollment was 28 years, most women (89%) were currently married, and fewer than half (40%) were employed. Fewer women living with HIV than HIV-uninfected women were married (84% vs 93%). Although more women living with HIV than HIV-uninfected women were employed (47% vs 34%), their median income was lower (10 vs 20 USD per week). Among women living with HIV, median time on ART was 5 years and median enrollment CD4 count was 547 cells/μl.

**Table 1.**
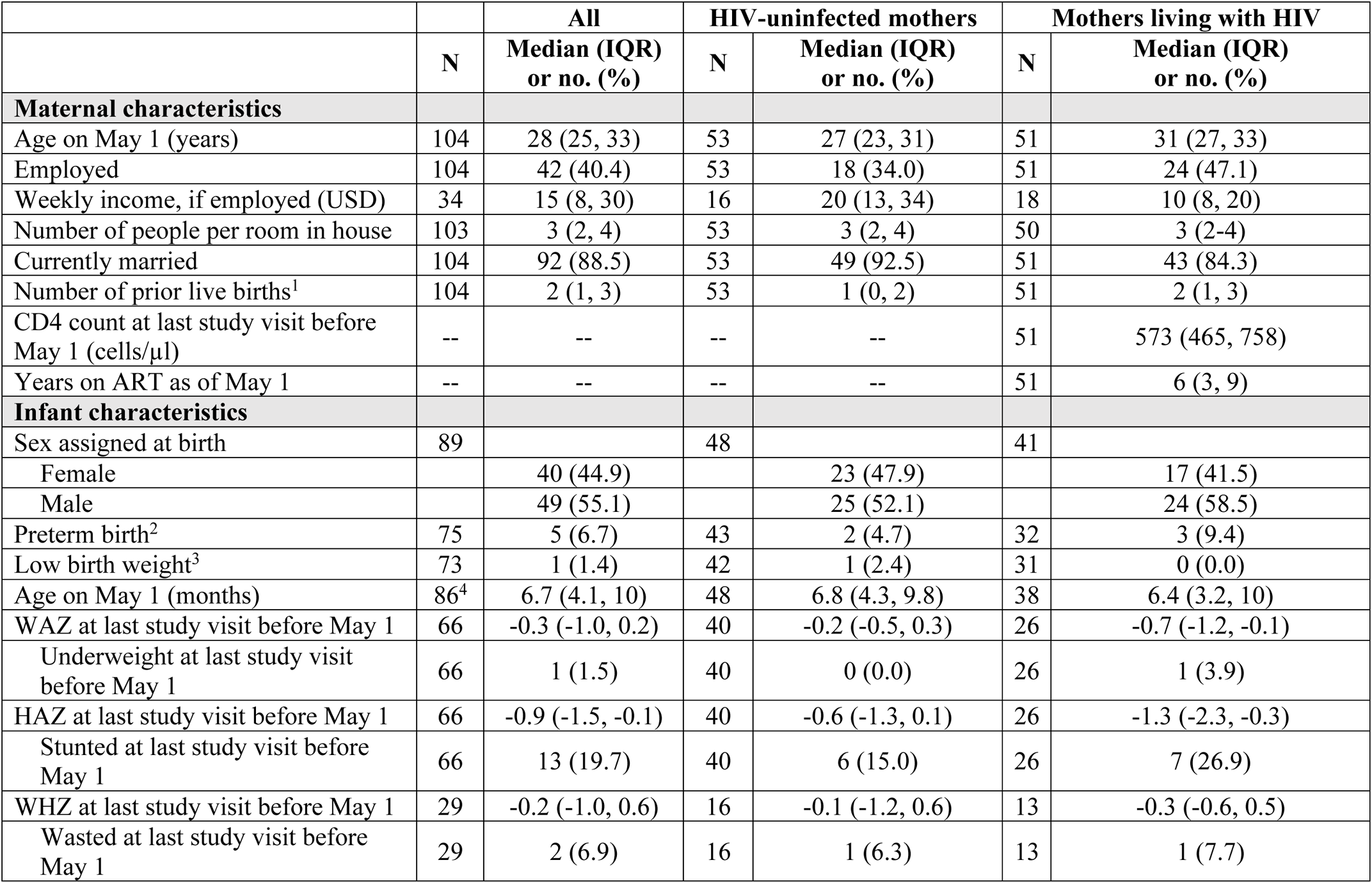
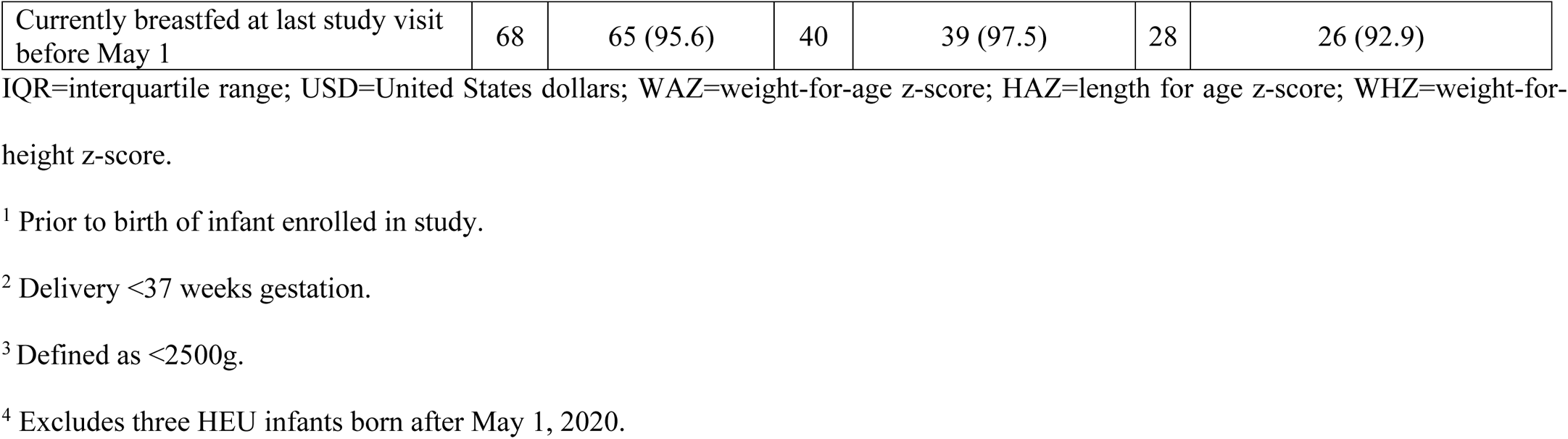
Participant characteristics.

Of the 89 infants, 55% were male, 7% were born preterm and 1% had low birth weight. The median age of infants on May 1, 2020 was 6.7 months. At their last study visit prior to May 1, 2020, nearly one-fifth (20%) of infants had stunting, 2% were underweight, and 7% had wasting; a greater proportion of HEU infants than HUU infants were stunted (27% vs 15%, respectively).

### Incidence of SARS-Co-V-2 Infection in Postpartum Women and their Infants

Thirty-nine cases of SARS-CoV-2 infection were identified among the 104 mothers (38%) and in 15 of the 89 infants (17%; Fig 1). Three of the mothers first tested positive before the start of the at-risk period and were excluded from analyses. All other cases first tested positive after May 1, 2020.

**Fig 1.**
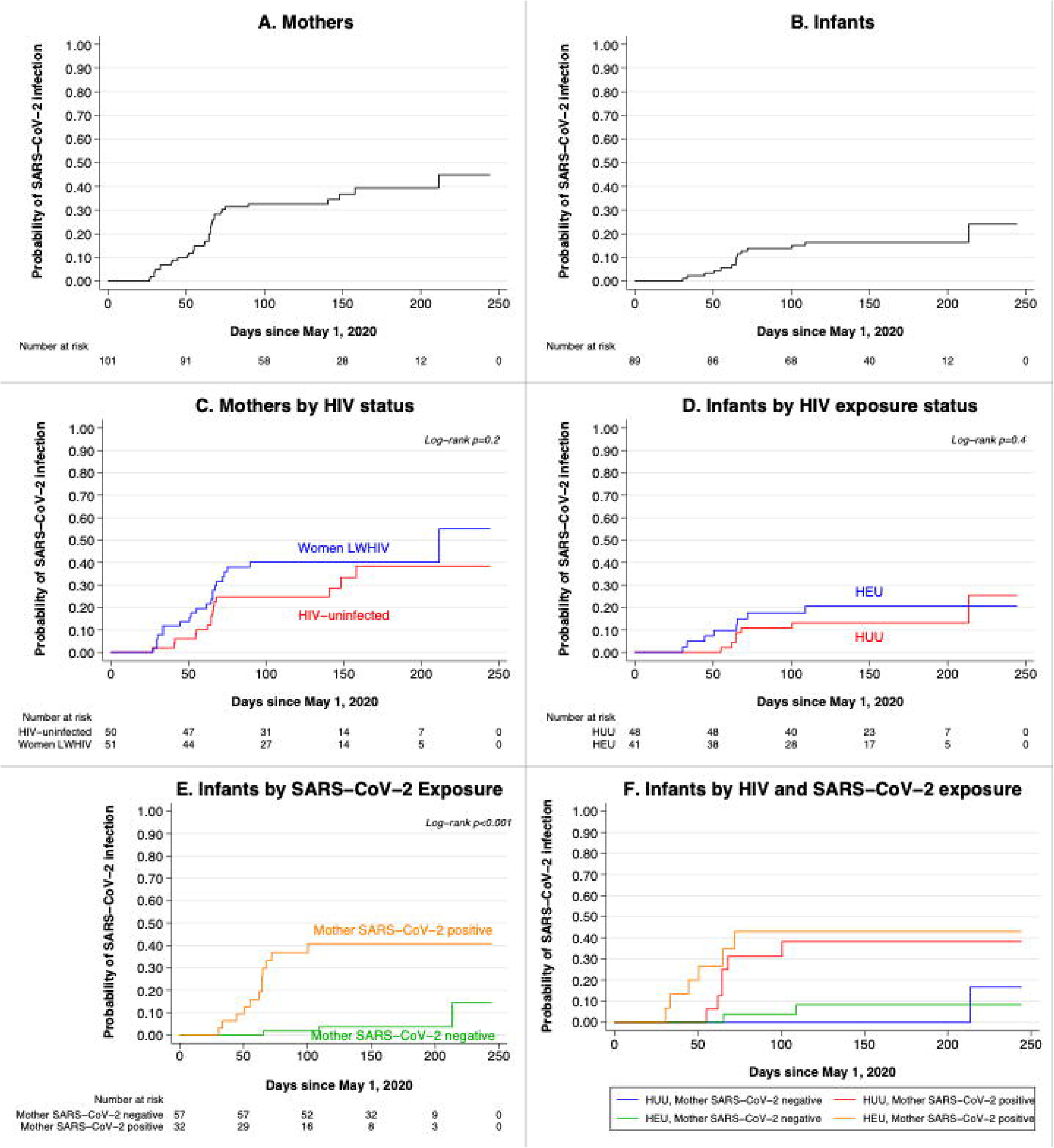
SARS-CoV-2 acquisition from May 1-December 31, 2020 in postpartum Kenyan women and their infants. Kaplan-Meier hazard functions for participants’ estimated date of infection are shown for (A) all mothers and (B) all infants, (C) mothers stratified by HIV status, (D) infants stratified by HIV exposure, (E) infants stratified by maternal SARS-CoV-2 infection, and (F) infants stratified by maternal HIV and SARS-CoV-2 infection. HEU=HIV-exposed uninfected, HUU=HIV-unexposed uninfected. All women enter the at-risk period on May 1, 2020; infants enter the risk period either at May 1 or on their date of birth, if after May 1. Timing of SARS-CoV-2 infection is estimated as the midpoint between the last negative and the first positive antibody test; for participants whose last negative antibody test was prior to May 1, timing of infection is estimated as the midpoint between May 1 and the first positive test.

The IR among individuals who tested positive after May 1, 2020 was 2.97 (95% CI: 2.14-4.12) per 1000 person-days among mothers and 1.20 (95% CI: 0.723-1.99) among infants (Table 2). The incidence of SARS-CoV-2 infection was 51% higher in women living with HIV compared to HIV-uninfected women and 48% higher in HEU versus HUU infants (Table 3), but these differences were not statistically significant (p=0.2 for mothers and p=0.4 for infants).

**Table 2.** Incidence of SARS-CoV-2 infection in postpartum Kenyan women and their infants from May 1 to December 31, 2020.

|  | Cases | Person-days<sup>1</sup> | Cases per 1000 person-days (95% CI) |
| --- | --- | --- | --- |
| **All mothers (N=101<sup>2</sup>)** | **36** | **12,116** | **2.97 (2.14, 4.12)** |
| HIV-uninfected (n=50) | 15 | 6370 | 2.35 (1.42, 3.91) |
| Living with HIV (n=51) | 21 | 5746 | 3.66 (2.38, 5.61) |
| **All infants (N=89)** | **15** | **12,482** | **1.20 (0.723, 1.99)** |
| **By HIV exposure** |  |  |  |
| HIV-unexposed (n=48) | 7 | 7096 | 0.986 (0.470, 2.07) |
| HIV-exposed (n=41) | 8 | 5386 | 1.49 (0.743, 2.97) |
| **By SARS-CoV-2 exposure** |  |  |  |
| Mother SARS-CoV-2 antibody-negative (n=57) | 3 | 8985 | 0.334 (0.108, 1.04) |
| Mother SARS-CoV-2 antibody-positive (n=32) | 12 | 3497 | 3.43 (1.95, 6.04) |

CI=confidence interval. HR=hazard ratio.
<sup>1</sup> Person time in days from May 1, 2020 or date of birth for infants born after May 1, 2020.
<sup>2</sup> Excludes three HIV-uninfected mothers who tested positive before May 1, 2020.

**Table 3.**
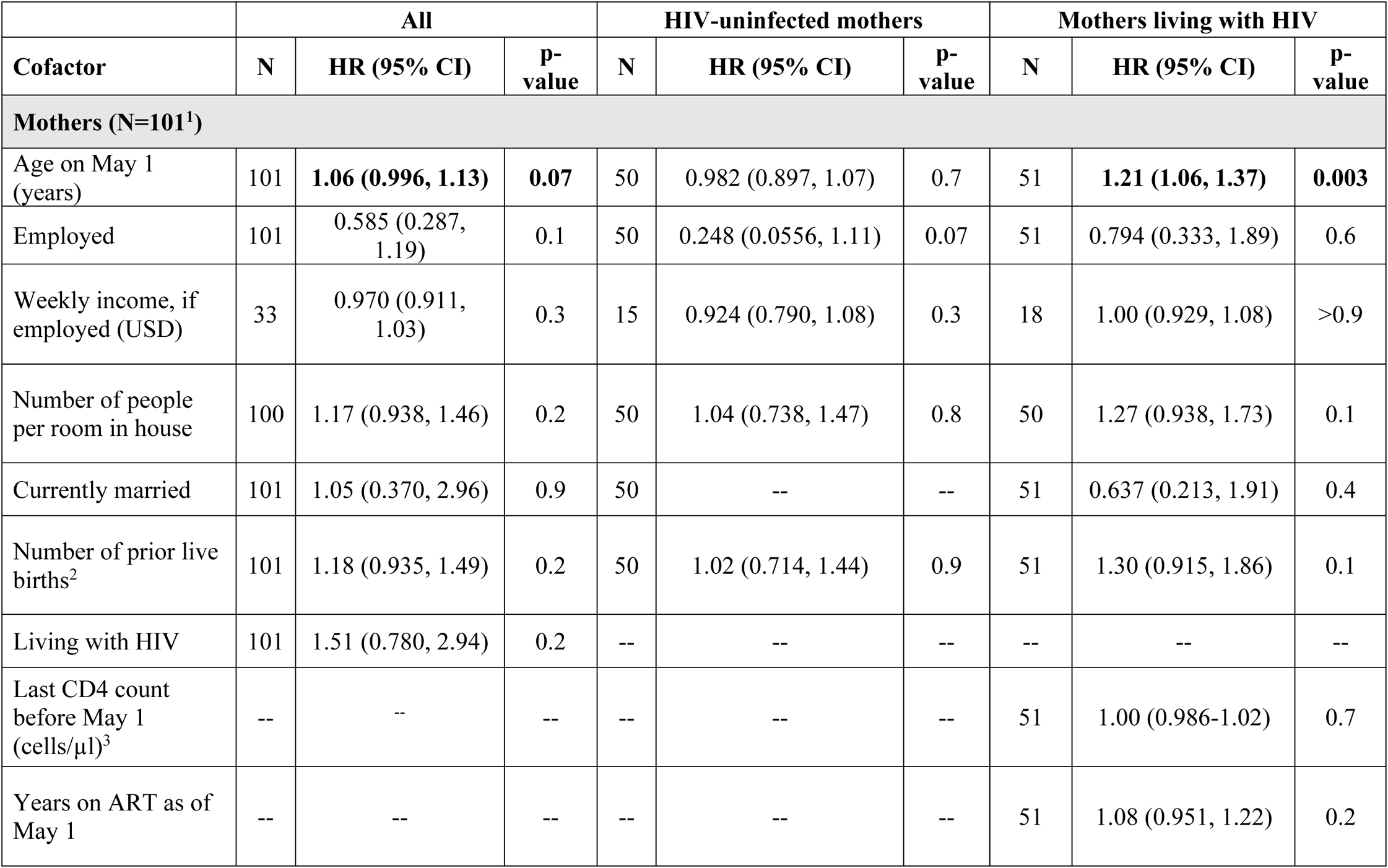
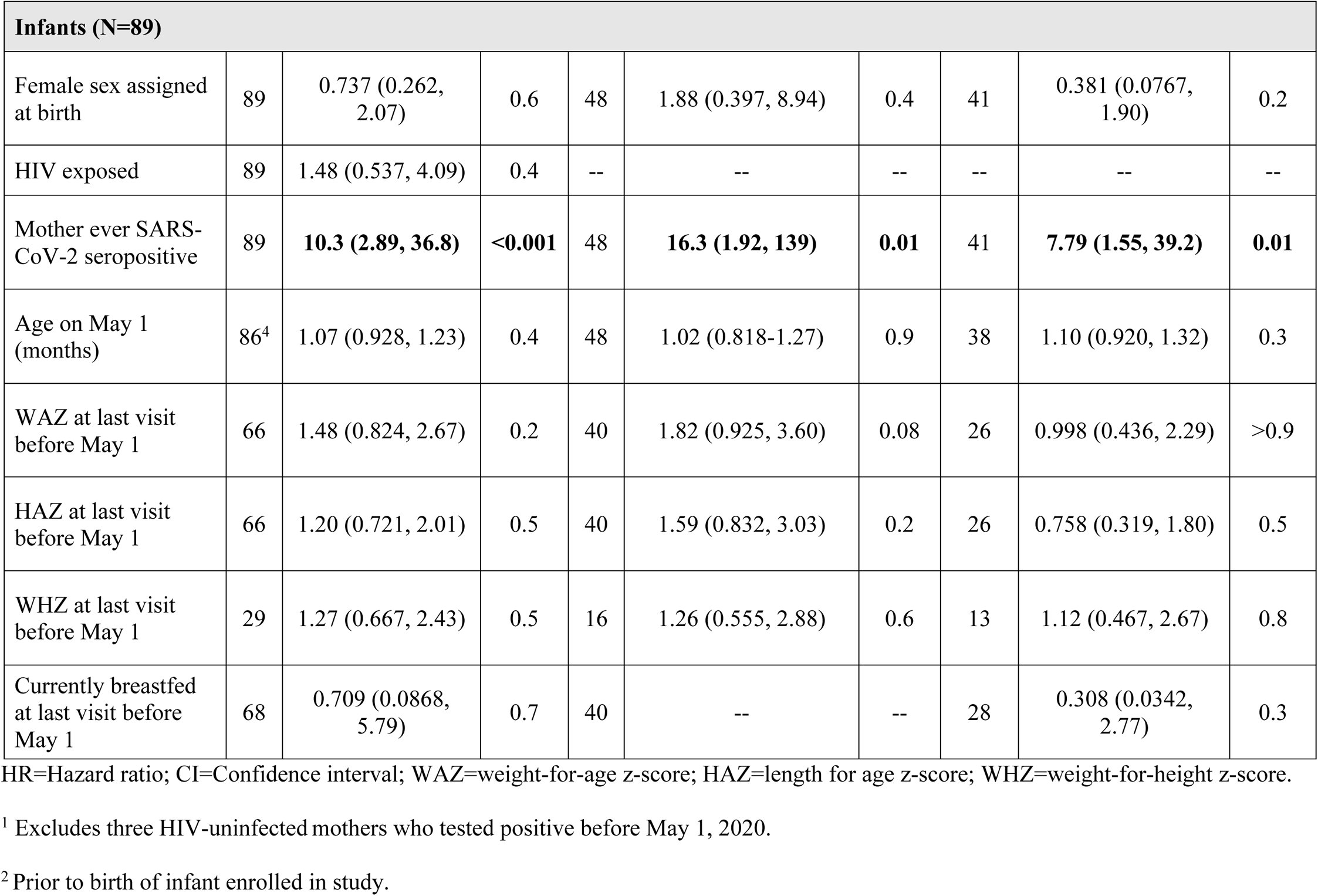
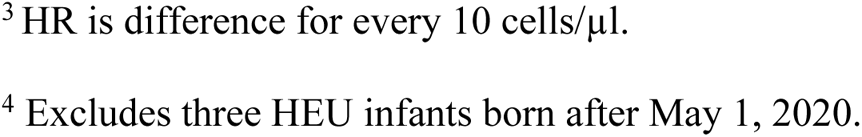
Correlates of SARS-CoV-2 infection in mothers and infants.

Infants whose mothers were SARS-CoV-2 seropositive were >10 times more likely to acquire infection (Hazard Ratio [HR]=10.3, p<0.001).

There was one SARS-CoV-2 seropositive infant for whom a false-positive test due to maternal antibody transfer during pregnancy could not be ruled out. The infant was born to a mother who tested negative for SARS-CoV-2 antibodies at enrollment at 28 weeks gestation but was seropositive at her first follow-up visit at 6 weeks postpartum. The infant’s first available blood sample, collected at week 10, was SARS-CoV-2 positive (OD ratio=1.66); subsequent samples were positive at month 6 (OD ratio=1.26) and negative at month 9 (OD ratio=0.371). Exclusion of this infant from analyses did not change incidence estimates or the hazard of infection associated with HIV or maternal SARS-CoV-2 infection (data not shown).

### Duration of SARS-CoV-2 Antibody Detection

We examined SARS-CoV-2 antibody changes over time in all participants with ≥1 test following their first positive (33 mothers and 13 infants). Antibody levels declined rapidly in both mothers and infants, regardless of HIV exposure, waning below the limit of detection in 9 (27%) women and in 4 (31%) infants (Fig 2A). The mean time between participants’ first positive serology test and loss of antibody detection was 5.6 months (95% CI: 4.9-6.4) for mothers and 4.8 (95% CI: 3.2-6.4) months for infants. Time to an antibody-negative test was not significantly different between women living with HIV and HIV-uninfected women (long-rank p=0.1) or HEU and HUU infants (p=0.2), though there was a trend for shorter mean time among women living with HIV and HEU infants (Fig 2B-C).

**Fig 2.**
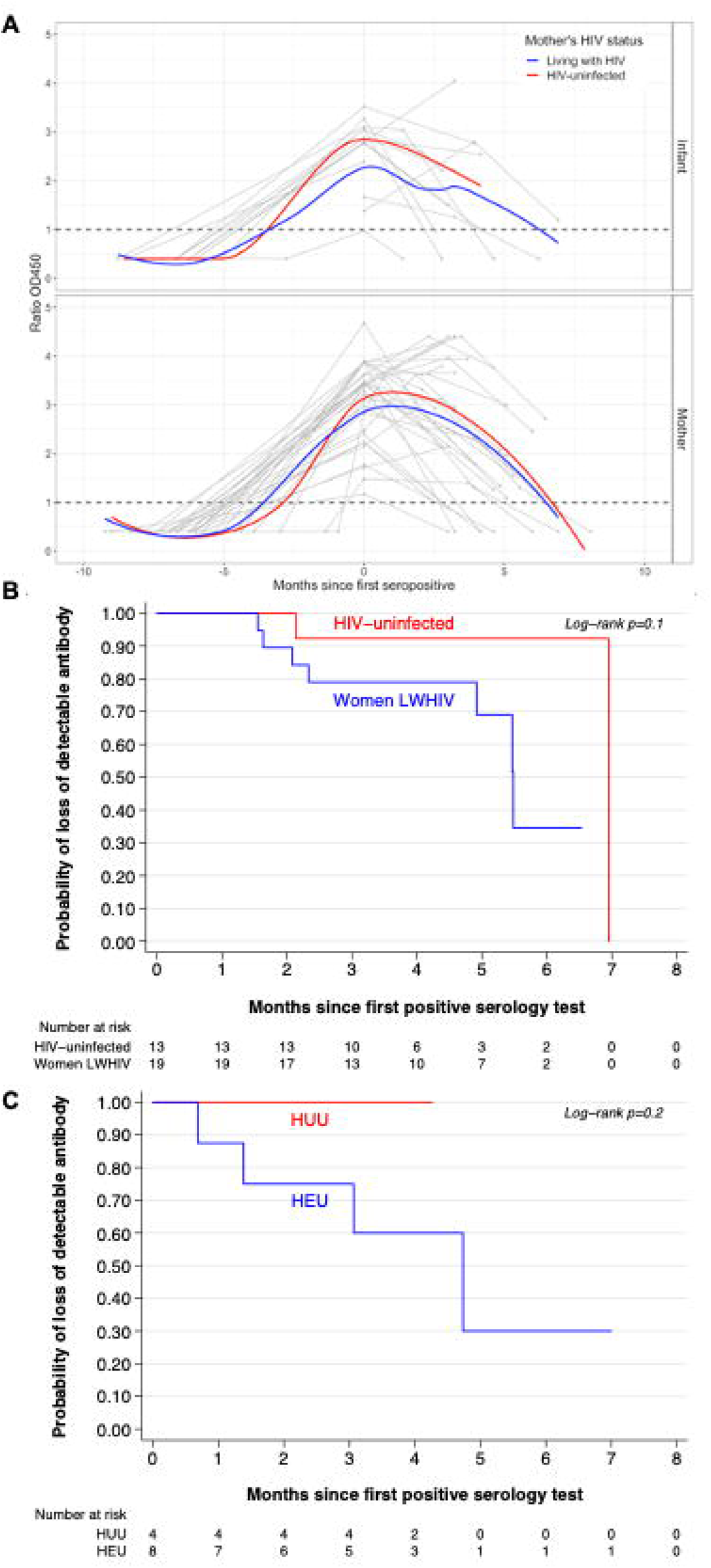
Detection of SARS-CoV-2 antibody among mothers and infants over time. (A) SARS-CoV-2 antibody levels over time relative to the first seropositive time point (0 months). Individual patterns in infants (top) and mothers (bottom) are shown in grey. Grouped by maternal HIV status, running means are shown for HIV-uninfected women or HIV-unexposed infants in black and women living with HIV or HIV-exposed infants in red. Limit of detection denoted by dashed vertical line. (B) and (C) are Kaplan-Meier hazard functions for participants’ estimated time to loss of detectable antibodies stratified by maternal HIV status and infant HIV exposure, respectively. HEU=HIV-exposed uninfected, HUU=HIV-unexposed uninfected. The risk period for loss of detectable antibody begins at the participant’s first positive serology test and ends either at the time of loss of detectable antibodies (estimated as the midpoint between the last positive test and first negative test after a positive test) or at the time of the most recent positive test.

Among 10 mother-infant pairs where both mother and infant were SARS-CoV-2 seropositive and had follow-up samples, 5 pairs remained concordant positive in all follow-up samples, 3 mothers lost antibody while their infant remained positive, one infant lost antibody while their mother remained positive, and in one case both mother and infant lost antibody concurrently (S1 Fig).

### SARS-CoV-2 Viral RNA Isolation from Stool

Twenty-seven mothers and 13 infants had stool samples collected between their last seronegative and first seropositive time points available for SARS-CoV-2 RNA testing. Of these, 5 mothers (19%) and one infant (8%) had detectable SARS-CoV-2 RNA (Fig 3). Two participants’ RNA-positive stool sample was collected on the same day as their first SARS-CoV-2 seropositive blood sample; the other 4 were RNA-positive prior to the first positive serology test. Median stool SARS-CoV-2 RNA level was 60,989 copies/ml (IQR 27,953-1,646,080) in mothers; the one infant’s RNA level was 10,181 copies/ml. SARS-CoV-2 genome sequencing was performed on six RNA-positive stool samples. High-quality whole genome sequences were obtained from two samples (S2 Fig). Both sequences were classified as B.1 lineage, a predominant global lineage that was also identified in Kenya in 2020.

**Fig 3.**
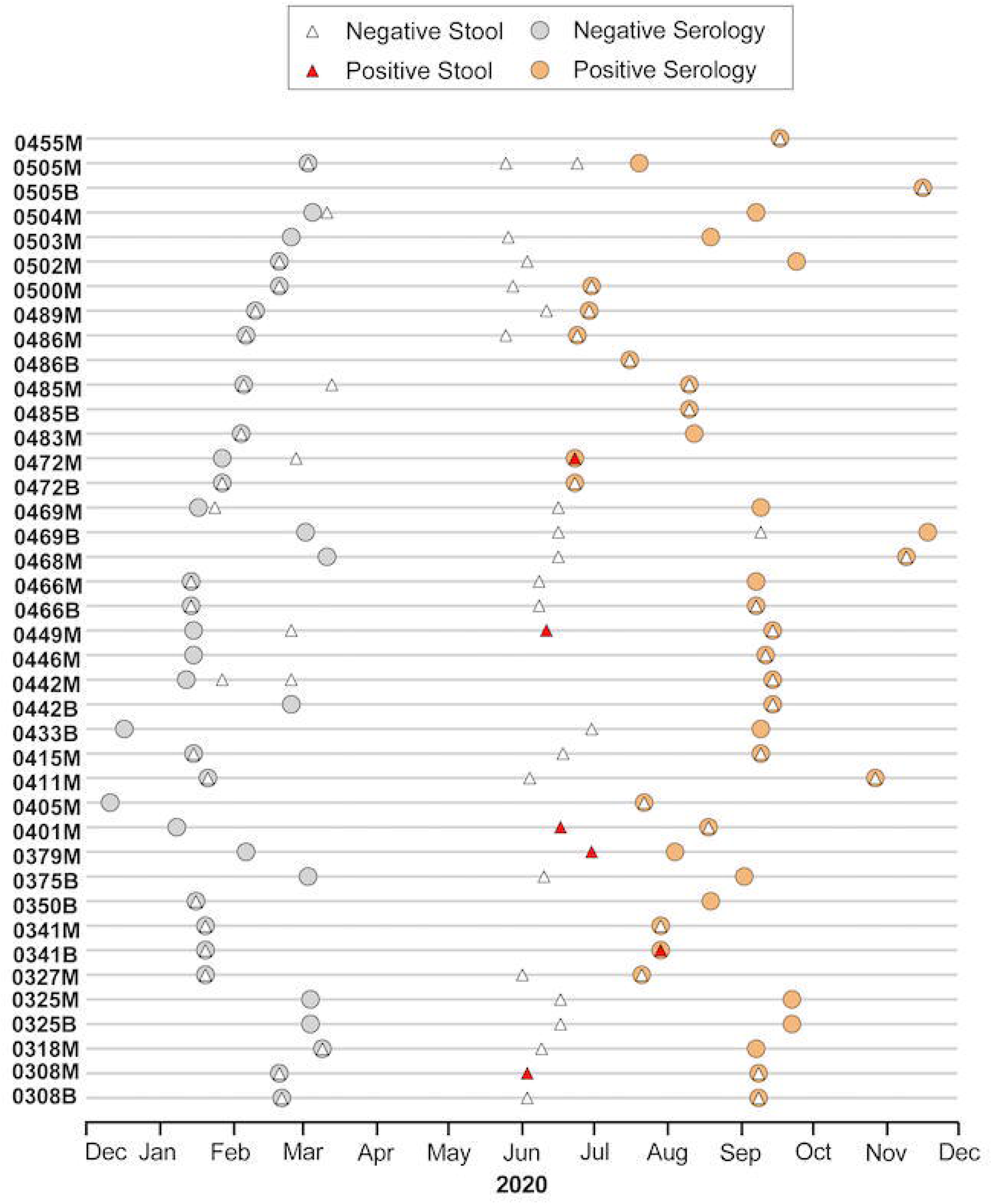
SARS-CoV-2 serology and stool viral RNA results over calendar time. Results of SARS-CoV-2 serology and quantitative real-time PCR testing of stool samples from Linda Kizazi participants that had a seropositive result between May 1-December 31, 2020. Anonymized ID numbers on y-axis for mothers (M) and infants (B). Grey circles indicate date of last seronegative serology test and orange circles indicate date of first seropositive sample. White triangles represent SARS-CoV-2 RNA-negative and red triangles represent RNA-positive stool samples. Calendar time is on the x-axis.

### Correlates of SARS-CoV-2 Infection

Older age was associated with increased risk of SARS-CoV-2 infection among women living with HIV (HR=1.22, 95% CI: 1.07-1.38) but not among HIV-uninfected women (HR=0.984, 95% CI: 0.900-1.08; Table 3). No other maternal characteristics were significantly associated with SARS-CoV-2 infection. Having a mother who was ever SARS-CoV-2 seropositive was significantly associated with an increased risk of SARS-CoV-2 infection for both HUU (HR=16.3, 95% CI: 1.92-139) and HEU (HR=7.79, 95% CI: 1.55-39.2) infants. No other infant characteristics were significantly associated with SARS-CoV-2 infection.

### Symptoms of SARS-CoV-2 Infection

There were no hospitalizations or deaths in the study cohort due to COVID-19. Approximately one-fifth of SARS-CoV-2 antibody-positive mothers (n=38, 19%) and infants (n=15, 21%) reported ≥1 symptom of COVID-19 at the time of their first seropositive visit or since their most recent visit prior. In mothers, symptoms were three times more likely at the first SARS-CoV-2 seropositive visit (RR=3.01, 95% CI: 1.43-6.45; p=0.005) compared to earlier seronegative visits. In infants, there was a trend to have a higher probability of having reported symptoms at their first seropositive visit compared to visits before SARS-CoV-2 infection (RR=1.93, 95% CI: 0.650-5.71; p=0.187).

## DISCUSSION

In this cohort of postpartum Kenyan women and their infants, 38% of mothers and 17% of infants experienced SARS-CoV-2 infection. Approximately 80% of infections were asymptomatic in both postpartum women and infants, and there were no cases of severe COVID-19 or death. In this study, we did not find a significant association between maternal HIV status and either maternal or infant SARS-CoV-2 infection risk. However, risk was dramatically increased among infants whose mothers acquired SARS-CoV-2 infection postpartum, indicating highly efficient intrahousehold transmission in this population. Seroreversion was commonly observed (27% and 36% of seropositive mothers and infants, respectively) during study follow-up. We detected shedding of virus in stool samples collected at and/or before seroconversion in 20% of adult participants, which confirmed our antibody results in these cases. Whole-genome sequencing of SARS-CoV-2 from stool recovered isolates that clustered with B.1 lineage variants, which were frequently identified both in eastern Africa and worldwide in 2020 [28–30]. Together, our data suggest that the initial two waves of COVID-19 in Kenya likely resulted in high rates of asymptomatic infection in postpartum women and their infants, and that detectable SARS-CoV-2 antibodies in these two populations waned rapidly.

This study is one of few that have examined the incidence of SARS-CoV-2 among postpartum women and their infants, including those living with HIV. Our findings among these women in living in a densely populated, urban neighborhood of Nairobi, are consistent with seroprevalence from a general population-based cross-sectional survey of households in November 2020, which found an age- and sex-adjusted seroprevalence of 34.7% in Nairobi County overall and 52.7% in the Mathare sub-county where the Linda Kizazi Study cohort resides [31]. Like our study, the survey noted a higher seroprevalence among adults (ages 20-59; 38.6%) than children (ages 0-9; 19.5%). Additionally, consistent with the rapid increase of COVID-19 in Kenya through the second half of 2020, we found a greater number of SARS-CoV-2 infections than a study of adult blood donors conducted early in the pandemic between April-June 2020, which showed just 7.8% of Nairobi County residents were seropositive [32]. However, our cohort’s incidence of SARS-CoV-2 infection was substantially higher than that of mother-infant pairs presenting with symptoms of COVID-19-like illness in a contemporaneous Siaya County, Kenya-based cohort study, which measured an incidence of 1.8 cases per 1000 person-months among postpartum women and 0.9 per 1000 person-months among infants [33]. This difference may be due to the high proportion of asymptomatic infections in Kenya.

While the incidence of infant SARS-CoV-2 infection was just under half that of mothers (17% vs 38%), infants whose mothers acquired SARS-CoV-2 infection were ten times more likely to become infected compared to infants not exposed to maternal SARS-CoV-2 infection. A recent meta-analysis of 54 intra-household transmission studies including >70,000 participants estimated a secondary attack rate of 17% overall, with rates higher in adults compared to children [34]. Though our analysis was unable to determine the household index case for participants in our study, the very high rate of infant infections and association with maternal SARS-CoV-2 infection suggest infant infections were likely acquired from their mother or a shared index case.

In this cohort, we did not find a significantly different risk of SARS-CoV-2 infection due to maternal HIV infection and CD4 count was not associated with risk of infection among women living with HIV. Although the differential risk of SARS-CoV-2 infection was 22% greater with each additional year of age among women living with HIV, this was not statistically significant.

Among mothers, the relative risk of symptoms was three-fold higher at the first seropositive visit compared to earlier visits. In contrast, seroconversion was not associated with symptoms of COVID-19 in infants, which is likely due to the high general incidence of mild infections experienced during the first two years of life, as well as the subclinical nature of many SARS-CoV-2 infections in young children [35]. There were no cases of severe COVID-19 or death among women in our cohort, which could be due in part to the young age of the participants. A study of the COVID-19 pandemic’s effects on children in sub-Saharan Africa (SSA) postulated that the larger populations of people <20 years in the region (52.7%) compared to Asia (31.2%), North America (24.5%), and Europe (21.2%) could partially explain the relatively low COVID-19 case burden and case fatality rate in this region [36]. However, it is also possible that limited surveillance and reporting in SSA underestimates the true burden of COVID-19 morbidity and mortality. Furthermore, the Linda Kizazi Study’s strict eligibility criteria generated a cohort of women that were mostly young, in good health, well-engaged in medical care, and, if living with HIV, had well-controlled HIV infection. All women in the analysis also delivered a healthy infant, another marker of maternal health. Thus, in this cohort of healthy postpartum women, we observed a lack significant impact of HIV co-infection and few symptoms overall, as compared to other populations living with HIV, in whom HIV has been associated with increased COVID-19 severity and death [10–15].

In seropositive mothers and infants with additional samples available post-SARS-CoV-2 infection, antibody levels waned over time and 30% had undetectable levels by a mean of 5-7 months. While there was no significant difference in time to loss of detectable antibody due to maternal HIV status or infant HIV exposure, there was a trend for shorter mean time to undetectable antibody among women living with HIV and HEU infants. Our data are consistent with earlier reports demonstrating mild and asymptomatic infections may have short-lived antibody detection [37–39]. Continued follow-up of participants during subsequent waves of Kenya’s epidemic will provide insights into both duration of antibody detection and the rates of reinfection.

Our study has several strengths, including prospective longitudinal testing of postpartum women living with HIV and HIV-uninfected women and their infants, and systematic and detailed assessment of clinical symptoms blind to COVID-19 infection status. We did not have viral RNA testing in nasal swabs to confirm serology results or detect acute SARS-CoV-2 infection, although we were able to verify the presence of virus in stool for some cases. Our study also has some important limitations to note, including a modest sample size, which may have precluded the detection of weaker associations. As discussed above, our selected population is not representative of the more heterogenous population of Kenyan postpartum women, women living with HIV, and infants due to our strict eligibility criteria. Most infants and mothers first tested SARS-CoV-2 antibody positive at the same visit, making it difficult to determine direction of infection, or to know whether both may have acquired infection from a shared index case. Three-month sampling intervals and missed visits during the start of the pandemic, while study procedures were being revised to be “no-contact” in accordance with Kenya Ministry of Health guidance, preclude precise ascertainment of timing of infection or detection of virus in most cases.

In summary, our data demonstrate high rates of asymptomatic and mildly symptomatic COVID-19 among healthy postpartum women with or without HIV co-infection during the initial waves of the pandemic in Kenya. Rapidly waning antibodies raise the possibility that despite high rates of infection, a large proportion of individuals may be susceptible to reinfection [40,41]. Women living with HIV and their infants were not found to be at a substantially increased risk of COVID-19 compared to HIV-uninfected women and their infants in this cohort, in contrast to other populations living with HIV. Continued practice of preventative measures such as social distancing and masking will remain important until COVID-19 vaccines become widely available in Kenya.

## Data Availability

The data required for this analysis were collected from human research participants and contain a set of sufficiently identifying variables, including dates of birth, that cannot be shared publicly per ethical guidance.

## ACKNOWLEDGEMENTS

We thank the Linda Kizazi Study participants, their families, and our study staff for their efforts to make this work possible. We thank the staff and leadership at Mathare North Health Centre and Kenyatta National Hospital in Nairobi, Kenya for supporting this research.

---

**S1 Fig. SARS-CoV-2 antibody levels over time in mother-infant pairs.** OD ratios show SARS-CoV-2 antibody levels over time in concordant SARS-CoV-2 seropositive mother-infant pairs with ≥1 sample available after initial antibody detection. Increased levels of antibody denoted by darker purple shading as shown in key. Positive antibody levels denoted by filled circle, equivocal levels by ⊗, and levels below the limit of detection by empty circles. Mother-infant pairs in which the mother was living with HIV are shown on top with bold red IDs; HIV-uninfected and -unexposed pairs are shown below with black IDs.

**S2 Fig. SARS-CoV-2 genomes sequenced from Kenyan stool samples.** Complete SARS-CoV-2 genomes were sequenced from six RNA-positive stool samples. (A) Phylogenetic analyses of 500 randomly selected SARS-CoV-2 global sequences, the Wuhan1 reference, and the two Kenyan stool-derived genomes (indicated in red) are shown. Clade labels are shown. (B) Next-generation sequencing data statistics of the six Kenyan stool samples.

