## Supplementary material for "HIV and SARS-CoV-2 infection in postpartum Kenyan women and their infants": S1 Appendix

***Detection of SARS-CoV-2 RNA in stool***

Stool samples (200mg) were diluted in 1200 µl of SMBuffer (G-Biosciences), then vortexed at 3,000rpm for two minutes. The homogenate was centrifuged at 7,000g for 10 minutes at 4°C and the supernatant was filtered through a 0.45µM filter. The filtrate was treated with 200µl of lysozyme/DNAse cocktail (100µl of Zero 10X Buffer (Lucigen), 4µl of Baseline-ZERO DNASE (Lucigen), 80µl of lysozyme (Sigma), and 16µl of SMBuffer) to enrich for virus-like particles for one hour at 37°C. Total nucleic acid was extracted using the bioMérieux eMAG instrument.

***SARS-CoV-2 RNA Sequencing***

SARS-CoV-2 genome sequencing was performed on samples that were SARS-CoV-2 RNA-positive. First-strand cDNA synthesis was performed using random hexamers (SuperScript III Reverse Transcriptase, Life Technologies), followed by PCR amplification of tiled amplicons spanning the SARS-CoV-2 genome (Swift Normalase Amplicon Panel, Swift Biosciences) and library construction. Libraries were sequenced on the Illumina NextSeq500 (v2.5, 2 x 150 paired end). Illumina sequencing reads were quality filtered to remove adaptors and low-quality bases using BBTools. High-quality-filtered reads were mapped to the SARS-CoV-2 Wuhan1 reference genome (NC_045512.2) using BWA-MEM (1) and amplicon primers were trimmed using Primerclip (version 0.3.8; available from https://github.com/swiftbiosciences/primerclip).

***Reference***

1. Li H. Aligning sequence reads, clone sequences and assembly contigs with BWA-MEM. 2013;1303.3997.
