## Supplementary figures and images for "HIV and SARS-CoV-2 infection in postpartum Kenyan women and their infants"

### S1 Fig

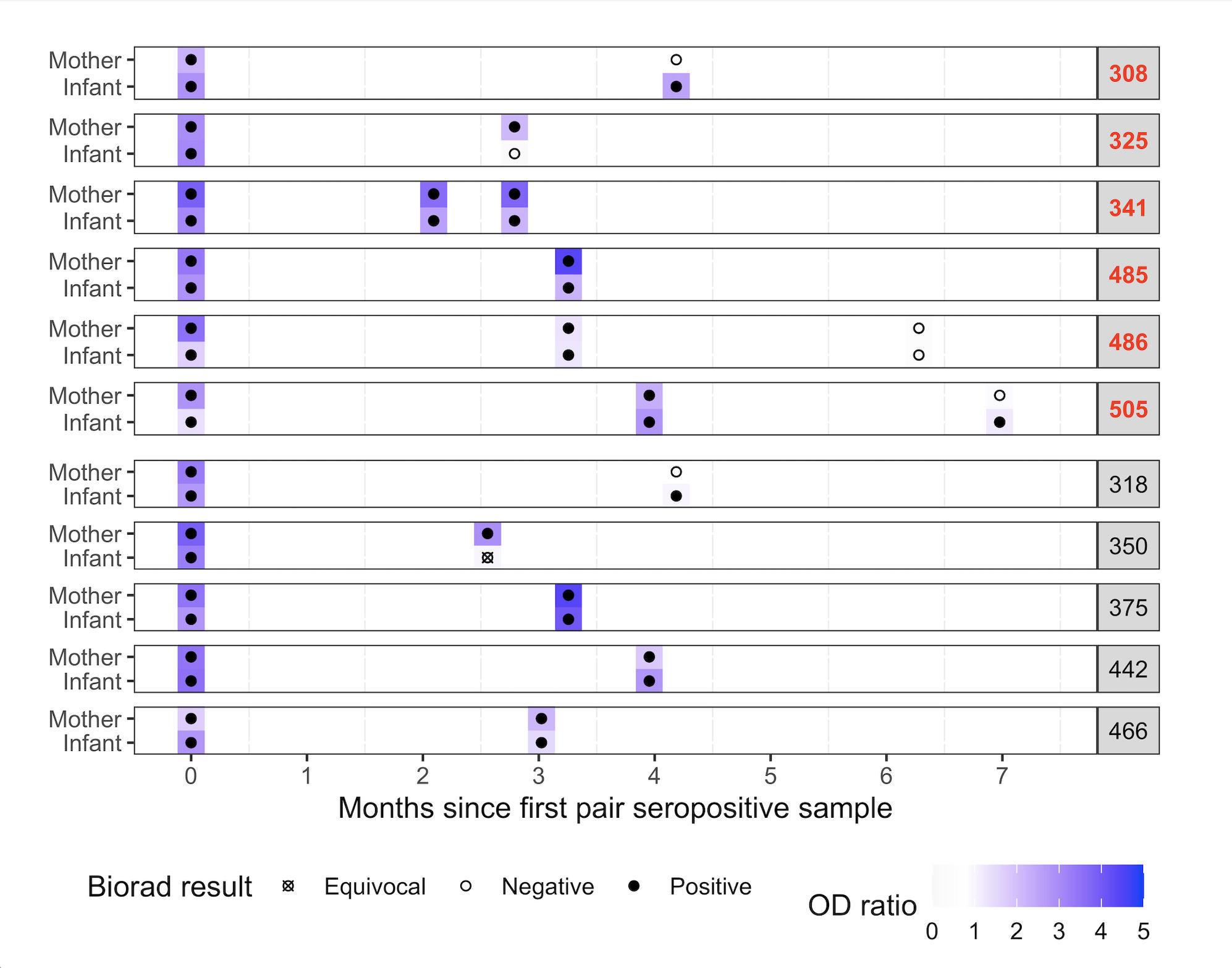

### S2 Fig

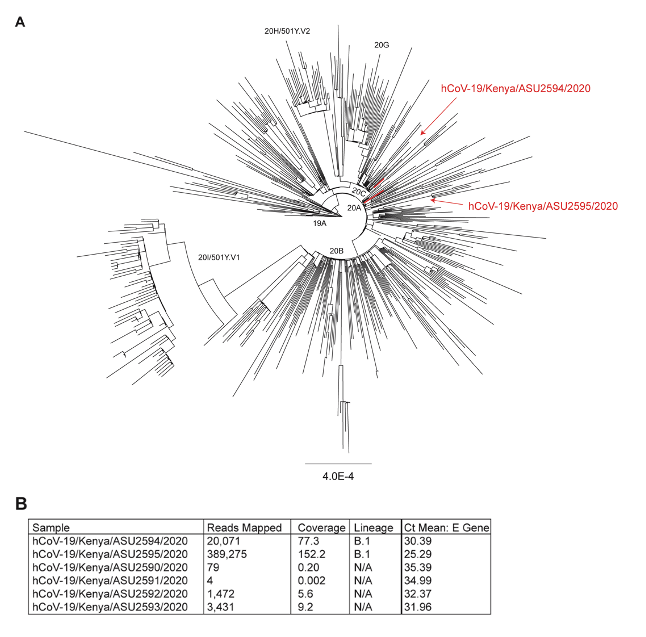
